# Primary healthcare providers’ perspectives on six-month dispensing of HIV medication in South Africa: a cross-sectional survey

**DOI:** 10.1101/2024.12.24.24319374

**Authors:** Vinolia Ntjikelane, Amy Huber, Allison Morgan, Sophie Pascoe, Musa Manganye, Lufuno Malala, Sydney Rosen

**Affiliations:** Health Economics and Epidemiology Research Office, Wits Health Consortium, Faculty of Health Sciences, University of the Witwatersrand, Johannesburg, South Africa; Department of Global Health, Boston University School of Public Health, Boston, MA, USA; HIV & AIDS Treatment, Care and Support Directorate, HIV & AIDS and STI Cluster, National Department of Health, Pretoria, South Africa

**Keywords:** 6MMD, antiretroviral therapy, 12-month prescriptions, healthcare providers, South Africa

## Abstract

**Background:** In many African countries the dispensing duration for antiretroviral therapy (ART) medication for HIV treatment has increased from 3 months to 6 months for stable clients. To help inform a decision about whether to move from three-month dispensing (3MMD) to six-month dispensing (6MMD) in South Africa, we surveyed healthcare providers about their perspectives on dispensing and scripting durations.

**Methods:** We conducted a cross-sectional survey of healthcare providers (nurses, managers, pharmacists) at 24 primary healthcare clinics in South Africa from May to September 2024, asking closed- and open-ended questions. Results are reported as frequencies.

**Results:** 182 providers were enrolled from four provinces (median age 44, 88% female). Most (>70%) respondents said that the 3MMD guideline offered multiple benefits for providers and patients, compared to the previous two-month dispensing rule; most (64%) also said there were no challenges in implementing 3MMD. >80% of respondents across all cadres reported that they would be comfortable dispensing 6 months of ART at a time, believing that it would reduce facility overcrowding, lighten staff workloads, and be advantageous to clients by decreasing their visit burden and travel costs. Two thirds (63%) of participating nurses, who provide the largest share of direct ART care, were also in favour of resuming 12-month scripting for ART; the remaining 37% expressed concerns about decreases in treatment adherence and clinical monitoring of clients.

**Conclusion:** Most healthcare providers at primary healthcare clinics in South Africa are in favour of allowing six-month dispensing and 12-month prescriptions as options for established ART clients.

## Background

Enrolment of established HIV treatment clients into differentiated models of care (abbreviated as DMOCs in South Africa; referred to as differentiated service delivery, or DSD, models in other countries) has been strongly recommended by the World Health Organization to increase the patient-centeredness of care and decongest healthcare facilities, while reducing provider and client costs and healthcare system burden(1). Multimonth dispensing of antiretroviral medications—at least a three months’ supply and as many as six months per healthcare system interaction--is a common component of many DMOCs(2). In many African countries, the number of months of medication provided to antiretroviral therapy (ART) clients at one time has increased from 1-3 months to 6 months(3–5). Under the six-month medication dispensing (6MMD) model, established clients typically visit the healthcare facility twice a year and receive a six-month supply of ART at each clinic visit. The goal of multi-month dispensing of ART is to lower patient-related barriers to treatment access, improve long-term retention in care, and lessen the burden of HIV treatment on healthcare systems, particularly in countries with limited resources, by reducing numbers of clinic visits required(6–9). Evaluations of 6MMD have found equal or slightly improved clinical outcomes such as viral suppression and retention in care for clients receiving six-month supplies, across multiple countries(4,5,8–11).

In South Africa, the recommended medication dispensing interval was increased in May 2023 from two to three months for established ART clients. South Africa’s DMOCs are supported by the Centralised Chronic Medicines Dispensing and Distribution (CCMDD) program (12,13) in most provinces; Central Dispensing Units and individual clinics provide pre-packaging of medications in other provinces and for ART clients not eligible for CCMDD. These programs enable clients to collect pre-packed medications from facility-based medication pickup points, external (community-based) medication pick-up points, and adherence clubs. These models intend to reduce barriers to care by reducing the frequency of facility visits, lessening time spent waiting in queues, and decreasing transport costs for clients (13,14). Established clients are issued six-month prescriptions and make a full clinic visit at least two times a year for clinical review and re-scripting. A three-month supply of medications is dispensed at each of these full clinic visits; between semi-annual clinic visits, clients pick up three-month supplies of medications at the pickup points.

Although South Africa’s 2023 ART guidelines do provide for six-month ART dispensing for established clients, depending on stock availability and operational capacity (15), 6MMD has not yet been rolled out nationally. South Africa’s National Department of Health (NDOH) is now considering whether to adopt six-month dispensing in the country and is examining questions that have been raised about supply chain limitations, facility storage needs, effect on client monitoring, and healthcare providers’ views on the desirability and feasibility of 6MMD. To help guide NDOH’s decision and understand the landscape for 6MMD in South Africa, we surveyed primary healthcare providers about their perspectives on longer ART dispensing and prescription durations, including six month dispensing and 12-month prescriptions.

## Methods

The SENTINEL study was a mixed-methods survey conducted at 24 primary healthcare facilities across four districts and provinces of South Africa: Alfred Nzo District in the Eastern Cape Province, Ehlanzeni in Mpumalanga, King Cetshwayo in KwaZulu Natal, and West Rand in Gauteng (16). Here we present results from the third round of SENTINEL data collection conducted from May 8 to September 18, 2024.

### Study sites, population, and data collection

The study protocol has previously been described in detail (16). Sites were selected for SENTINEL to provide diversity in setting (rural v urban) and capture the variation in DMOCs offered in South Africa at the time of study launch (2021). For this analysis, we asked the facility manager at each study site to identify potentially eligible healthcare providers for the survey. Providers were eligible if they were directly or indirectly involved in ART and DMOC implementation, were employed in their roles at their respective study sites for at least six months, and provided written informed consent to participate in the study. We aimed to enrol up to ten providers at each site, with a sample size based on study resources and the size of the relevant staff complement at most study facilities.

Trained research staff explained to the eligible providers why the study was being conducted and informed them that participation was voluntary and that they could stop the interview at any point. Research staff then conducted the informed consent process and administered the survey in person to the providers in a confidential room at the study site, at a time that was convenient to the provider. Interviews were conducted with, among other staff, facility managers, nurses at different professional levels, and pharmacists and pharmacy assistants.

Using a primarily quantitative instrument designed for this study, data were collected about providers’ characteristics and roles, their perspectives regarding the benefits and challenges of the current 3-month ART dispensing duration and 6-month prescription length, and their opinions about six multi-month ART dispensing durations and 12-month scripting. We also included qualitative, open-ended questions to allow providers to expand upon their perspectives and provide their own opinions on why each dispensing or prescription duration would be acceptable or unacceptable.

During the COVID-19 pandemic, the maximum ART prescription duration allowed was increased from 6 to 12 months and required clinic visits reduced to one per year, to facilitate uninterrupted ART adherence during the lockdown period and to reduce over-crowding and limit interpersonal contact (13,14,17). Permission for 12-month scripting was not renewed, and South Africa returned to the usual 6-month prescriptions in September 2021. We thus also asked participants their views on returning to 12-month scripting.

### Data analysis

For this analysis, we limited the analytic data set to facility managers, nurses, and pharmacists and pharmacy assistants, all of whom interact directly with clients as clinical professionals. We first produced descriptive statistics to summarise participants’ characteristics, including their role and employer (government Department of Health or a nongovernmental partner). We then analyzed responses to quantitative questions about their perspectives on current ART dispensing durations and their views on longer ART dispensing and prescription durations. We report these as frequencies and percentages.

Open-ended responses were analysed using conventional qualitative content analysis methods(18) in Microsoft Excel. The coder, trained in qualitative content analysis, used an inductive approach to coding responses, identifying themes as they emerged. Similar themes were merged as the qualitative coder deemed appropriate. Mentions of emergent themes were tabulated and most common themes are described. Illustrative quotes were selected to represent emergent themes and are provided with staff cadre of the respondent.

### Ethical considerations

Ethical approval for SENTINEL was granted by University of Witwatersrand (Medical) Human Research Ethics Committee (Protocol M210241) and the Boston University Medical Campus IRB (H-41402). The protocol was also approved by the South African Provincial Health and Research Committees through the National Health Research Databases for each study province and district. All enrolled healthcare providers provided written informed consent for participation in the study. The parent study (SENTINEL) is registered as NCT05886530.

## Results

### Respondent characteristics

We enrolled in the survey a total of 149 facility managers, nurses and pharmacists, and pharmacist assistants whose responses were included in this analysis. Participants had a median age of 45 years and were predominantly female (90%), as shown in Table 1.

**Table 1.** Provider characteristics stratified by provider role (N=149)

| Characteristic | Facility managers | Nurses | Pharmacists and pharmacy assistants | Total |
| --- | --- | --- | --- | --- |
| N | 15 | 124 | 10 | 149 |
| Age in years (median, IQR) | 53 (49,57) | 44 (36,52) | 39 (33,49) | 45 (37,53) |
| Years in current role (median, IQR) | 7 (4,10) | 5 (3,10) | 10 (4,11) | 6 (3,10) |
| Gender (female) | 14 (93%) | 113 (91%) | 7 (70%) | 134 (90%) |
| Employer |  |  |  |  |
| Department of Health (government) | 15 (100%) | 118 (95%) | 9 (90%) | 142 (95%) |
| Nongovernmental partner | 0 (0%) | 6 (5%) | 1 (10%) | 7 (5%) |
| Employment status |  |  |  |  |
| Full time | 15 (100%) | 116 (94%) | 8 (80%) | 139 (93%) |
| Contract | 0 (0%) | 8 (6%) | 2 (20%) | 10 (7%) |

### Benefits and challenges of three-month ART dispensing

Nearly all providers (N=147, 99%) reported that the current maximum ART dispensing duration in their respective clinics was 3 months. The remaining providers (N=2, 1%) indicated that the maximum dispensing duration was 2 months, reflecting the previous guidelines before May 2023.

Providers described many benefits and few challenges in dispensing 3 months of ART to clients. They said that in comparison to 1- or 2-month dispensing, 3-month dispensing decreased clinic visits (87%), decreased facility workloads (80%), was convenient to clients (76%), and reduced clinic waiting times for clients (71%) (Figure 1). Nearly two thirds (64%) of the providers reported that there were no challenges encountered with dispensing 3 months of ART to clients. For those who mentioned challenges, these included medication stockouts (15%), difficulty in clinical monitoring of clients (13%), and concerns about client adherence (11%). No providers mentioned facility storage of a larger volume of medications or inventory loss as a concern.

**Figure 1.**
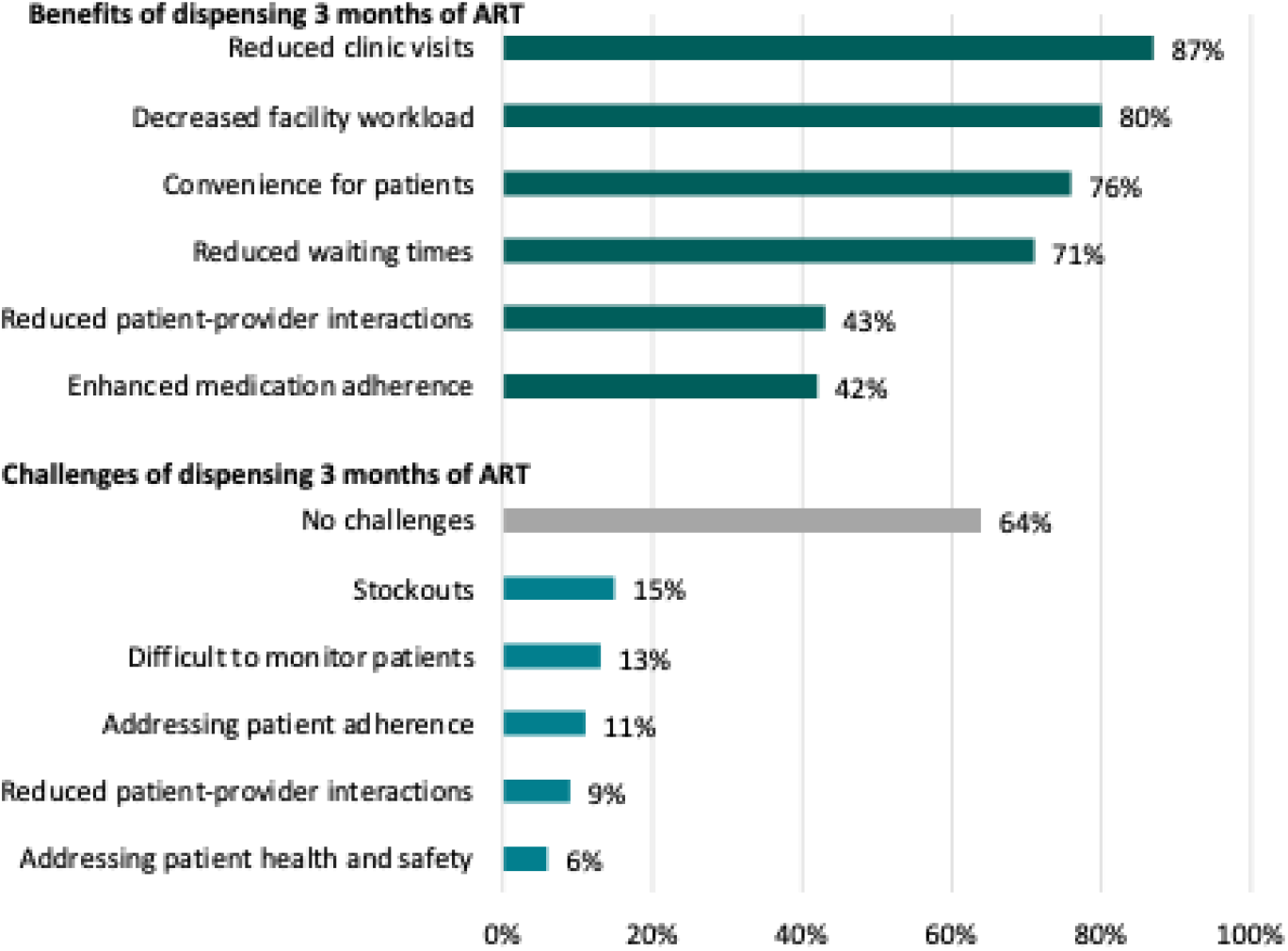
Provider reported benefits and challenges of 3-month ART dispensing (N=149)

### Providers’ views on six-month ART dispensing

We found that 60% of providers surveyed would like to see changes in dispensing intervals, beyond the current three-month limit. A larger majority, 84%, said they would be comfortable dispensing 6 months of ART; this response was most common among nurses (85%) (Figure 2). Providers perceived that 6MMD would make their work easier, by decongesting facilities and decreasing their workloads and the pressure they felt. They also stated that in their views, 6MMD would be appropriate for clients who are established on ART and that it would reduce clients’ travel burden and cost of seeking care.

**Figure 2.**
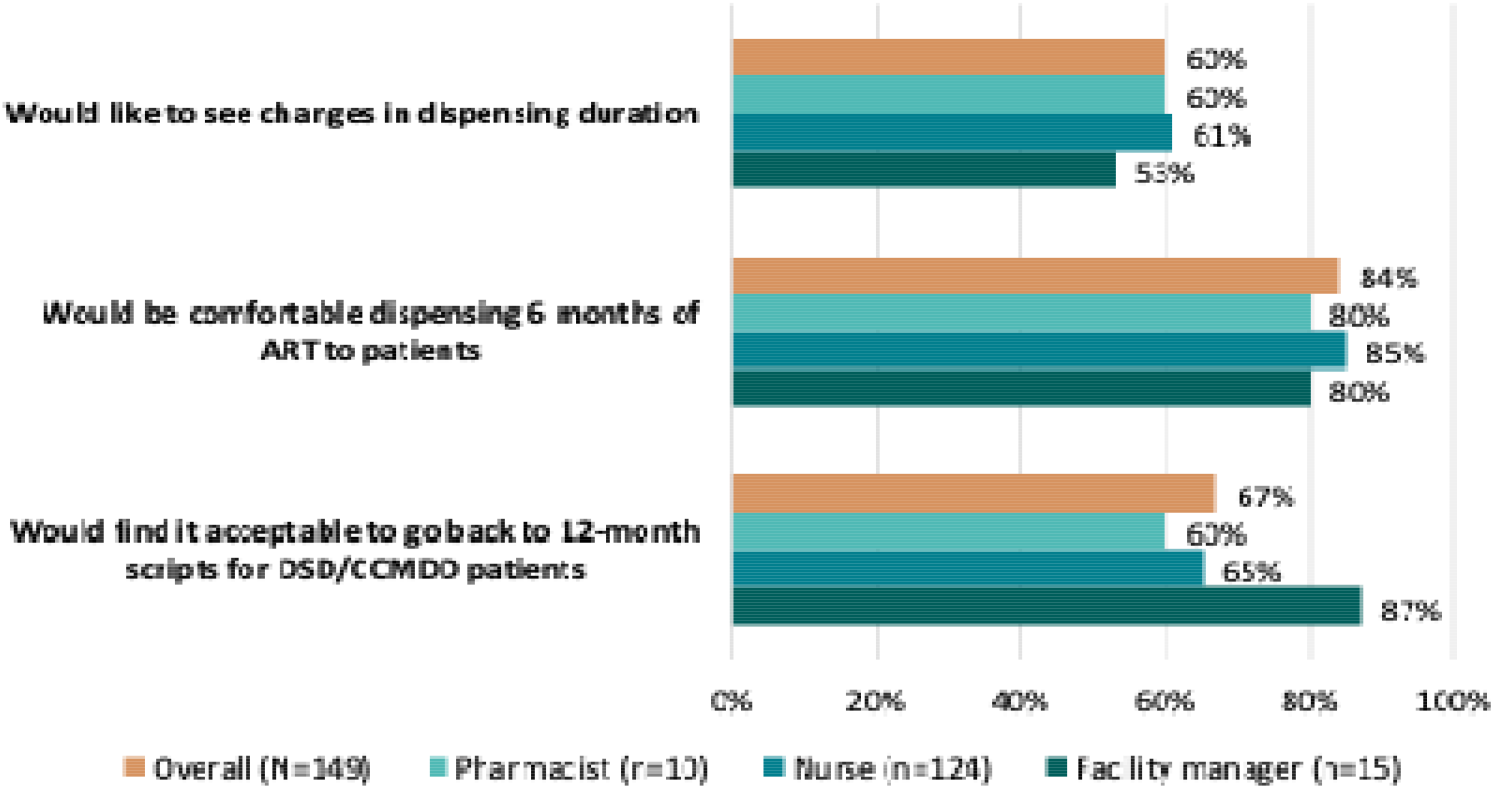
Providers’ views on longer ART dispensing durations, stratified by provider role.

Qualitatively, providers who said they were comfortable with 6MMD (125/149, 84%) stated that an increase in dispensing intervals would be ideal for working clients and would reduce facility workloads.

> *“Dispensing duration [should] be extended to at least 6 months which will reduce the frequency of medication refills and allow for more efficient management of clients” – Professional nurse*
>
> *“6 months duration would be ideal as clients have work commitments.” – Professional nurse*
>
> *“It would make life to easier for clients who are stable and we can be able to reduce the work load at the facility and we can give more attention to other clients who need urgent attention. – Professional nurse*
>
> *“It reduces the workload at the facility and also help clients not to use much money and time on transport fare” – Staff nurse*

A minority of providers (24/149, 16%) said they would not be comfortable with 6MMD and raised concerns regarding adherence and retention if 6-month dispensing were to be offered.

> *“We won’t be able to manage clients properly. We will be having a lot of challenges in adherence*.*” – Professional nurse*
>
> *“People do not honor their visit dates they would be lost follow up” – Pharmacist assistant*

Figure 2 also reveals that a high proportion (87%) of the facility managers and 65% of nurses said that they would find it acceptable to return to 12-month scripting, as was permitted during the COVID-19 pandemic. Similar to their perceptions of 6MMD, respondents said that 12-month prescriptions would benefit both clients and providers by improving workload, reducing crowding, and requiring fewer client interactions. Other providers mentioned that longer scripting durations will empower clients by encouraging self-management of their health.

> *“Firstly, we won’t experience congestion in the facility. Secondly, we are experiencing staff shortage in this facility and by having few clients will mean less stress for us. Thirdly, it will reduce the number of clinic visits for our clients who often find it hard to adhere to appointment dates*.*” – Facility manager*
>
> *“This will encourage clients to self-manage. Empower them to take charge of their health and be able to contact providers should they have any problems” – Registered Nurse*

The one third of providers who found going back to 12-month scripting unacceptable were concerned about decreases in adherence and clinical monitoring of clients.

> *“I feel that it would be difficult to monitoring clients who receive scripts for 12 months. It would be very risky… it will increase the number of clients who default on medication” – Professional Nurse*
>
> *“Being away too long will cause havoc. Not all Clients are reliable” – Professional Nurse*

For all outcomes reported above, we saw no meaningful difference in mean age, number of years of experience at the study site, or rural v. urban setting between those in favour of longer dispensing and/or scripting and those expressing concerns about these changes,

## Discussion

In this survey of South African ART providers, more than 80% of respondents across all cadres reported that they would be comfortable dispensing 6 months of ART at a time, believing that it would reduce overcrowding in the facilities and staff workloads and be advantageous to clients by decreasing their visit burden and travel costs. For the current maximum dispensing duration for ART of three months—an increase from the earlier maximum of two months--providers pointed to many benefits and few challenges. Two thirds of participating nurses, who provide the largest share of direct ART care, were also in favour of resuming 12-month scripting for ART. A large majority of respondents said that the three-month dispensing guideline recently adopted offered multiple benefits for providers and patients, compared to the previous two-month dispensing rule.

Our findings are consistent with the few others on this topic reported in the literature. In Ethiopia, providers and clients reported positive experiences with 6MMD, including perceived reduction in client time and costs, improved quality of care, and healthcare system benefits such as reduced provider workload and decongested facilities(19). Six-month dispensing has a largely favourable record for clinical outcomes(11), as noted above, and most clients, including adherence club participants in South Africa(20,21), have reported that they prefer it to shorter dispensing durations, though some wish to retain more frequent clinic contact(19,22). In a recent qualitative study in South Africa, providers preferred 12-month prescriptions due to the independence it gave clients and reduced number of clients seen in facilities, though, as in our survey, some providers were concerned about adherence to treatment and clinical monitoring of clients (13).

Should South Africa decide to offer stable ART clients six-month dispensing for HIV treatment, a number of relatively modest changes will be needed, mainly at the facility level, to accommodate it. These could include, for example, modifying package sizing and labelling, ensuring adequate inventory and storage space, and adjustment of current counselling messages to incorporate the option of receiving a six-month supply at a time. Use of facility resources (staff time and clinic space) is not likely to change substantially, since existing DMOCs already require only two clinical consultations per year, with intermediate medication supplies provided at medication pickup points. As mentioned above, moreover, procedures for 6MMD are already included in national guidelines. Providers’ concerns about not being able adequately to monitor and manage patient welfare should also be addressed, through, for example, early analysis of outcomes data to determine if specific subgroups of patients do less well with 6MMD than with the previous 3MMD. Adapting treatment literacy and counselling messages to reflect clients’ responsibilities and potential needs between six-month dispensing intervals may also help address providers’ concerns. In contrast to six-month dispensing, twelve-month prescribing would require a national policy change and thus may be a longer-term goal. Capacity assessment of a sample of healthcare facilities is currently underway to provide additional information to the NDOH as it makes its decision.

This study provides some of the first evidence of healthcare providers’ perspectives on six-month dispensing and 12-month prescriptions in South Africa. Respondents represented multiple cadres of providers from four different provinces and diverse facilities, and the inclusion of both quantitative and qualitative questions provided a comprehensive picture of provider views. The study does have several limitations, however. Participants were drawn from only 24 of South Africa’s more than 3,000 primary healthcare facilities and may thus not be representative of the country as a whole. Views on 6MMD and 12-month dispensing reflected individuals’ expectations of what such procedures would entail, rather than actual experience under the specific guidelines that are adopted. As with other survey results, our responses might also reflect a degree of social desirability bias or, in contrast, status quo bias, leading participants to express more or less enthusiasm for the innovation proposed (e.g. 6MMD) than they truly feel.

## Conclusions

As South Africa’s National Department of Health considers the pros and cons of adopting a national policy allowing six-month dispensing of antiretroviral medications for HIV, this study suggests that ART providers would welcome such a change, believe it will be beneficial to providers and patients, and do not foresee problems with it. Some providers express concerns about poorer ability to manage and monitor patients, and these issues should be considered in determining who should be eligible for 6MMD and how to respond to HIV-related health concerns that arise between six-month clinic visits. It is likely, however, that 6MMD would not differ greatly from South Africa’s current DMOC approach with regard to monitoring and management and appears to offer a promising strategy for reducing clinic congestion and providers job burdens, improving patient experiences (time, cost), and, in general, making ART more patient-centred.

## Acknowledgements

The authors would like to thank all the healthcare providers who took part in the study and the AMBIT SENTINEL research assistant team for conducting the interviews.

## Author contributions

SP, AH, VN, and SR conceptualized and designed the study. AH and VN supported the implementation of the study. VN oversaw and supervised data collection. VN, AH, and AM led data analysis and data curation. VN drafted the manuscript. MM and LM interpreted the results. All authors reviewed, edited and approved the final draft of the report.

## Competing interests

The authors report no conflicts of interest. MM and LM are employees of the government agency that has oversight authority for the study sites.

## Ethics

Ethical approval for SENTINEL was granted by University of Witwatersrand (Medical) Human Research Ethics Committee (Protocol M210241) and the Boston University Medical Campus IRB (H-41402). The protocol was also approved by the South African Provincial Health and Research Committees through the National Health Research Databases for each study province and district. All enrolled healthcare providers provided written informed consent for participation in the study.

## Funding

This research was funded by the Gates Foundation under INV-037138 to the Wits Health Consortium. The funder had no role in study design, data collection and analysis, decision to publish, or preparation of the manuscript.

## Data availability

Disidentified data will be posted in a public repository following closure of the study protocol with the supervising ethics committees; the corresponding author can be contacted for details.

## Notes

### Competing Interest Statement

The authors have declared no competing interest.

### Clinical Protocols

https://bmchealthservres.biomedcentral.com/articles/10.1186/s12913-023-09813-w

